# Ukrainian syndrome. Behavioral and functional changes in Ukrainian youth under the conditions of a 2022-Full-scale Russian invasion

**DOI:** 10.1101/2023.01.08.23284307

**Authors:** Anatolii Tolstoukhov, Mykhailo Matiash, Spartak Subbota, Vitalii Lunov

## Abstract

The Ukrainian syndrome is a certain behavioral pattern of adaptation and an individual position of a Ukrainian regarding the Russian-Ukrainian war. In this context, we proceeded from the model of the spatial-relational disposition of a traumatic event, which reflects the degree of objectivity of the loss / threat at the individual, family-related levels, and the level of the immediate environment (Lunov, 2022).

In the conditions of a sharp increase in mental and behavioral disorders caused by the Russian invasion, the system of providing mental care and departmental psychological services turned out to be such, at least in the first stages, that the established models of assessment and intervention need to be revised (Mosiichuk, Tkach, Lunov, 2022).

One of the reasons for this is that until today there are no generalizations in Ukraine regarding national norms of mental health and psychological well-being of young people. There is also a lack of research on the perception and understanding of the behavioral, functional, and psychosomatic changes experienced by young people under the influence of the prolonged Russian-Ukrainian war.

The purpose of our research is to determine the behavioral and psychosomatic changes and risks perceived by Ukrainian youth due to the prolonged Russian-Ukrainian war, in the time perspective after the full-scale Russian invasion on February 24, 2022.

**Materials and methods:** The “Scale of non-clinical assessment of behavioral changes and social functioning” (block A, B. Rate on a scale: -1 means this sign used to be characteristic of me, but now it has disappeared, or it does not bother me; 0 means I practically do not feel it, or it is absent; 1 means there are certain signs, but they do not significantly affect life and well-being; 2 means the manifestation of this sign is strong; 3 means I feel exhausted and disorganized), and the Questionnaire of the experience of being in a combat zone developed by Vitalii Lunov were used in the study. The study was conducted from April 2022 to January 2023.

4694 youth and young scientists aged 16 to 40 from 61 cities of Ukraine took part in the research.

The conducted research demonstrates the generalization of descriptive statistics of behavioral, functional, and psychosomatic changes experienced by Ukrainian youth in the conditions of the protracted Russian-Ukrainian war.

According to the obtained results, it can be stated that the following behavioral and functional patterns of Ukrainian youth require the attention of psychologists:

- behavior characterized by increased fatigue, exhaustion, weakening or loss of ability to prolonged physical or mental stress, irritability, frequent mood swings, tearfulness, moodiness, vegetative disorders.
- behavior characterized by subjective experiences (feelings of anxiety, own inferiority, fear of heights, closed spaces, obsessive thoughts, memories, etc.), somatic and vegetative disorders (disturbed sleep, poor appetite, vomiting, diarrhea, etc.).
- indecisiveness and tendency to excessive reasoning, alarming incredulity, ease of forming obsessive fears, apprehensions, actions, rituals, thoughts, ideas.
- decrease in vital activity.
- mental health problems.
- significant changes in habitual behavior.

We remind you that the obtained results are based on the respondents’ self-reports. However, the most significant thing for us is the youth’s self-awareness of their behavioral, psychosomatic, and functional changes.

Such awareness becomes the basis for understanding the new norm of life during the war.

## Introduction

As we noted in our previous studies, “Ukrainian syndrome” is the result of any mental trauma: war, natural disaster, accident, mutilation, violence (sexual or domestic) (Matiash, Khudenko, 2014). The Ukrainian syndrome is a certain behavioral pattern of adaptation and an individual position of a Ukrainian regarding the Russian-Ukrainian war. In this context, we proceeded from the model of the spatial-relational disposition of a traumatic event, which reflects the degree of objectivity of the loss / threat at the individual, family-related levels, and the level of the immediate environment (Lunov, 2022).

The results of our previous studies of the psychological health of Ukrainian youth reveal its criterial content and adaptive-resource conditionality. In this context, we note the heuristic potential of the paradigms of humanistic and behavioral psychology regarding healthy functioning according to the principle of balance-imbalance, as one of the values of the sociogenesis of Ukrainian youth (Lunov, Lytvynenko, Maltsev, Zlatova, 2022).

**The purpose of our research** is to determine the behavioral, functional, and psychosomatic changes and risks perceived by Ukrainian youth due to the prolonged Russian-Ukrainian war, in the time perspective after the full-scale Russian invasion on February 24, 2022.

## Materials and methods

The “Scale of non-clinical assessment of behavioral changes and social functioning” (block C, D. Rate on a scale: -1 - this sign used to be characteristic of me, but now it has disappeared, or it does not bother me; 0 - I practically do not feel it, or it is absent; 1 - there are certain signs, but they do not significantly affect life and well-being; 2 – the manifestation of this sign is strong; 3 – I feel exhausted and disorganized), and the “Questionnaire of the experience of being in a combat zone” developed by Vitalii Lunov were used in the study. The study was conducted from April 2022 to January 2023.

The research protocol was approved by the Department of General and Medical Psychology of the Bogomolets National Medical University (protocol No. 15 dated 31.03.2022). The survey of respondents was conducted on the platform https://forms.gle/U99akzToALh2mXLc9. All respondents gave their consent to use the results of the research for scientific purposes.

4694 youth and young scientists aged 16 to 40 from 61 cities of Ukraine took part in the research.

The research was carried out in accordance with the scientific topics and projects of the laboratory of the psychology of learning of the G.S. Kostyuk Institute of Psychology of the National Academy of Educational Sciences of Ukraine “The potential of genetic psychology in the study of the interaction of subjects in the educational space” (No. 0121U107603, 2021-2023), the department of general and medical psychology of the Bogomolets National Medical University “Methodological, clinical, applied aspects of psychological medicine and psychological practice” (No. 0120U100656, 2020-2022). The research was carried out with the support of the grant of the LLC “National Institute of Evidence-Based Psychotherapy” “Metacognitive psychology and evidence-based psychotherapy” (No. 2022-MPEBP, 2022-2024).

## Results and Discussion

Considering the behavioral changes of Ukrainian youth in the conditions of the Russian-Ukrainian war and the full-scale Russian invasion, the following was established.

According to the results of the self-assessment by Ukrainian youth, “***Behavior characterized by increased fatigue, exhaustion, weakening or loss of ability to prolonged physical or mental tension, irritability, frequent mood changes, tearfulness, moodiness, vegetative disorders***” was recognized.

3.8% of respondents believe that this feature used to be characteristic of them, but now it has disappeared or does not bother them. 17.3% of respondents say that fatigue, exhaustion, weakening or loss of ability to long-term physical or mental stress, irritability, frequent mood swings, tearfulness, moodiness, autonomic disorders are practically not felt by them, or are absent.

At the same time, 30.0% of respondents testify that there are certain signs of such behavioral changes, but they do not significantly affect life and well-being. At the same time, 32.7% of respondents say that they feel certain symptoms, but they do not significantly affect their life and well-being. However, 16.2% of Ukrainian youth admit it. That the manifestations of such behavioral changes are strong, and 16.2% feel exhausted and disorganized in connection with this.

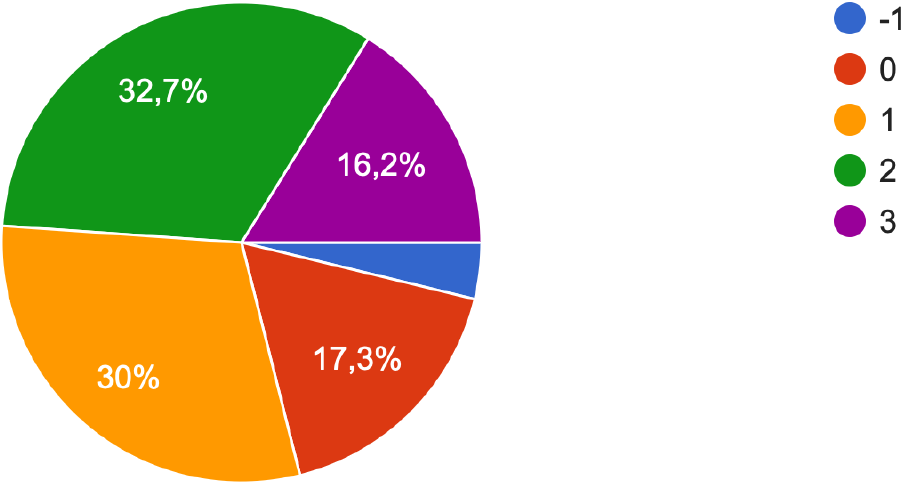

The next question was self-assessment “***Behavior characterized by feelings of anxiety, own inferiority, fear of heights, closed spaces, obsessive thoughts, memories, etc***., ***somatic and vegetative disorders (disturbed sleep, poor appetite, vomiting, diarrhea, etc.)***”.

In the study, it was established that only 3.7% of the respondents indicated that this feature used to be characteristic of them, but now it has disappeared or does not bother them. 23.9% of respondents practically do not feel the specified behavioral property. However, 32.9% noted that there are certain signs of such behavior, but they do not significantly affect life and well-being.

At the same time, which requires special attention, 28.9% of respondents noted that the manifestation of such behavior is strong, and 10.6% feel exhausted and disorganized due to the inability to master such behavior.

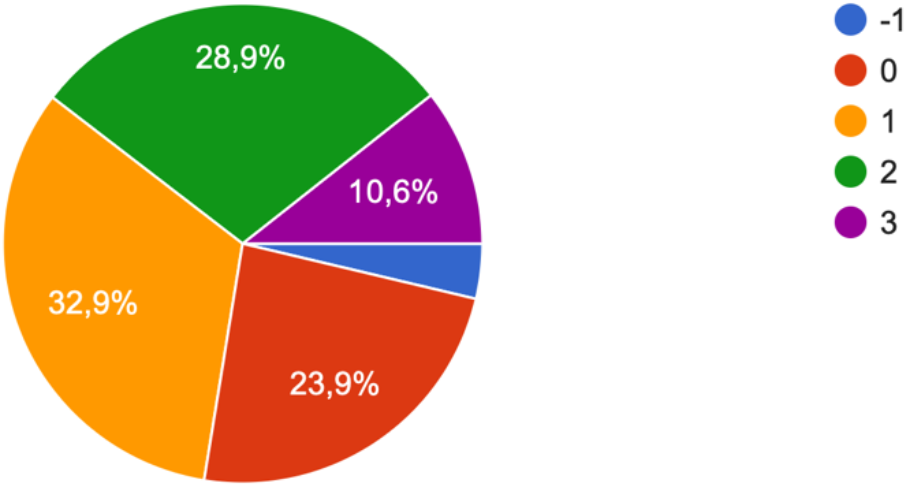

Evaluating one’s own behavior according to the indicators “***Boundless egocentrism, insatiable thirst for constant attention, admiration, surprise, respect, compassion. Lying and fantasizing are completely aimed at embellishing one’s persona. Absence of deep sincere feelings due to great expression of emotions, theatricality, tendency to posturing***” the respondents gave the following answers.

Thus, in the opinion of 7.1% of respondents, this sign of behavior was characteristic of them in the morning, but now it has disappeared or does not bother them. 68.6% of respondents practically do not feel its influence on behavior.

At the same time, 16.2% of respondents found that there are certain signs of it, but they do not significantly affect life and well-being, and 6.3% testified that the manifestation of this sign is strong, and 1.8% feel exhausted and disorganized because of this form of behavior.

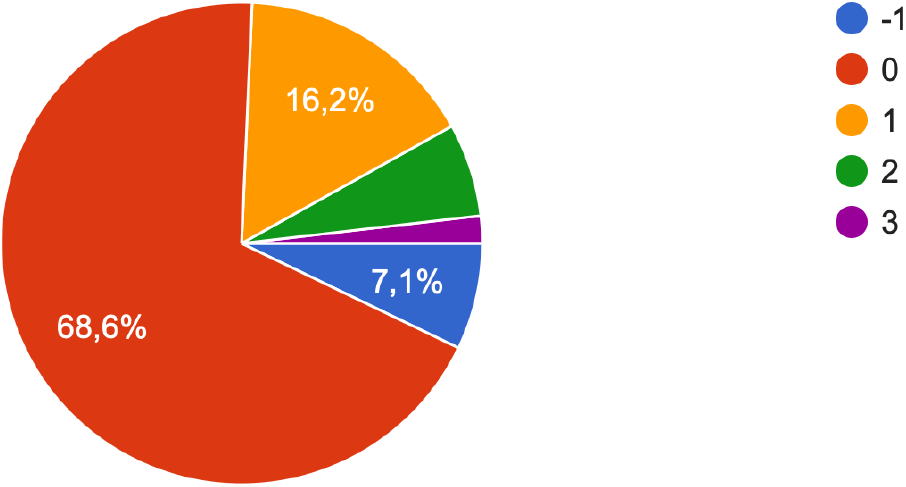

According to the results of the self-assessment of such behavioral manifestations in Ukrainian youth as: “***Indecision and tendency to excessive reasoning, anxious distrust, ease of forming obsessive fears, apprehensions, actions, rituals, thoughts, ideas***” the following results are summarized.

5.6% of respondents believe that this feature used to be characteristic of them, but now it has disappeared or does not bother them. 33.7% of respondents stated that they practically do not feel such changes in their behavior.

At the same time, 32.1% say that there are certain signs of the mentioned behavior, but they do not significantly affect life and well-being. It is noteworthy that 23.1% of respondents state that the manifestation of such behavior is strong, while 6.0% feel exhausted and disorganized due to such behavioral changes.

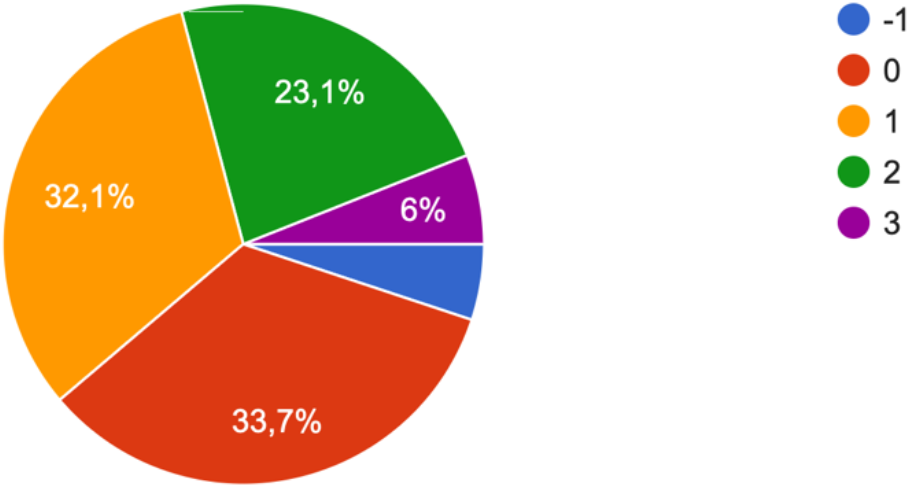

Regarding the assessment of “***Features of behavior associated with “bad character”, protest reactions, antisocial behavior***”, the respondents noted the following.

4.8% of respondents noted that this behavior used to be characteristic of them, but now it has disappeared or does not bother them. 58.9% of respondents noted that they practically do not experience such behavioral changes, or they are absent.

At the same time, 25.3% of respondents determined that there are such signs in their behavior, but they do not significantly affect their life and well-being. However, 9.1% of respondents claim that the manifestation of such behavioral changes is strong; and 1.9% feel exhausted and disorganized due to such changes in behavior.

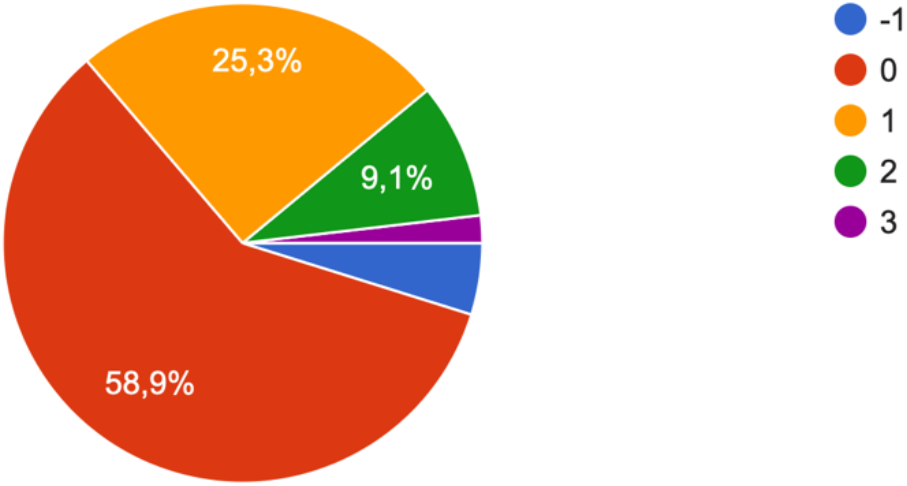

Assessing behavior with ***symptoms of cerebral origin (dizziness, psychosensory disorders, etc.)***, respondents demonstrated the following.

Thus, 4.4% of respondents say that such symptoms used to be characteristic of them, but now they have disappeared or do not bother them. 71.6% of respondents practically do not experience such symptomatic changes.

17.0% say that there are certain signs of the specified symptoms, but they do not significantly affect life and well-being. In addition, 5.6% claim that the manifestation of this feature is strong, and 1.4% feel exhausted and disorganized.

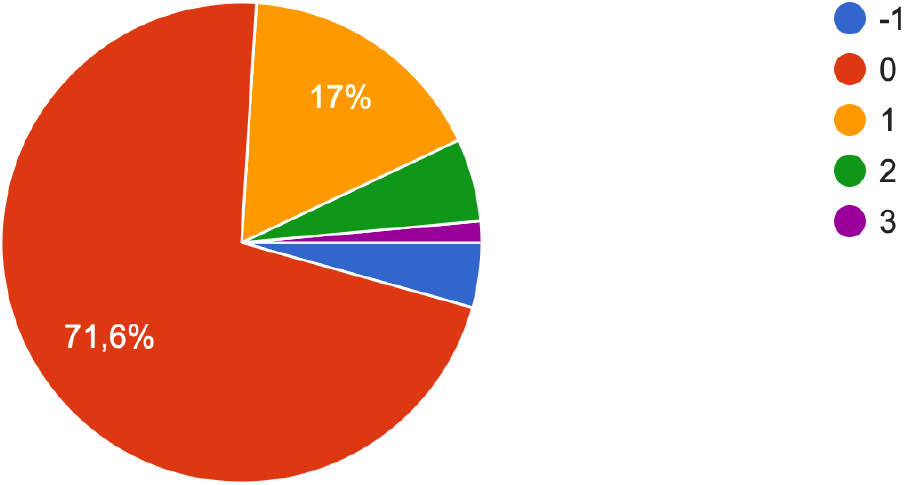

It should also be noted about the respondents’ self-assessment of ***symptoms related to the functioning of the ear-throat-nose system***.

According to 5.6% of respondents, this feature used to be characteristic of them, but now it has disappeared or does not bother them. 61.3% of respondents do not feel their influence, or it is absent. While 23.5% of respondents testify that there are certain signs, however, they do not significantly affect life and well-being. It should be noted that 8.5% of respondents say that the manifestation of this feature is strong, and 1.1% feel exhausted and disorganized.

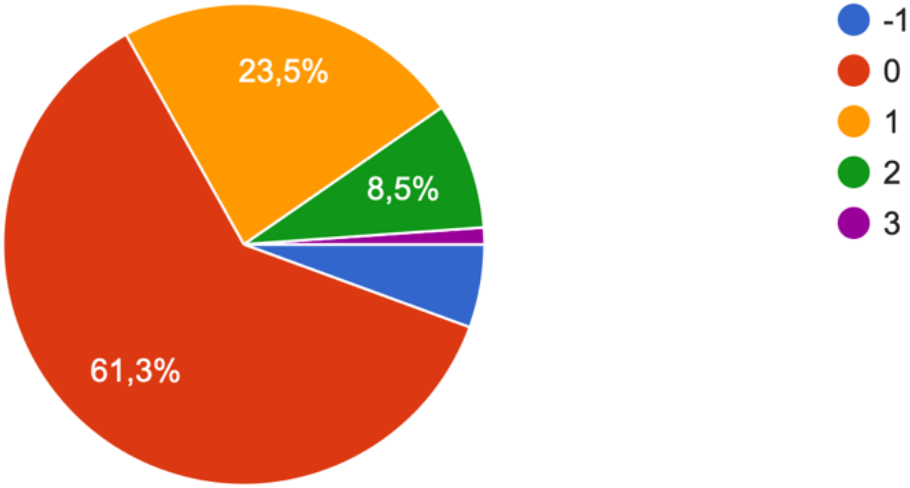

Evaluating the ***symptom complex reflecting the deterioration of the gastrointestinal functional system*** in Ukrainian youth, the following was established.

5.7% of respondents found that this symptom complex used to be characteristic of them, but now it has disappeared or does not bother them. 48.3% practically do not experience these symptoms. 29.4% noted that there are certain signs of these symptoms, but they do not significantly affect life and well-being.

At the same time, for 14.1% of respondents, the manifestation of this symptom is strong, and 2.5% feel exhausted and disorganized.

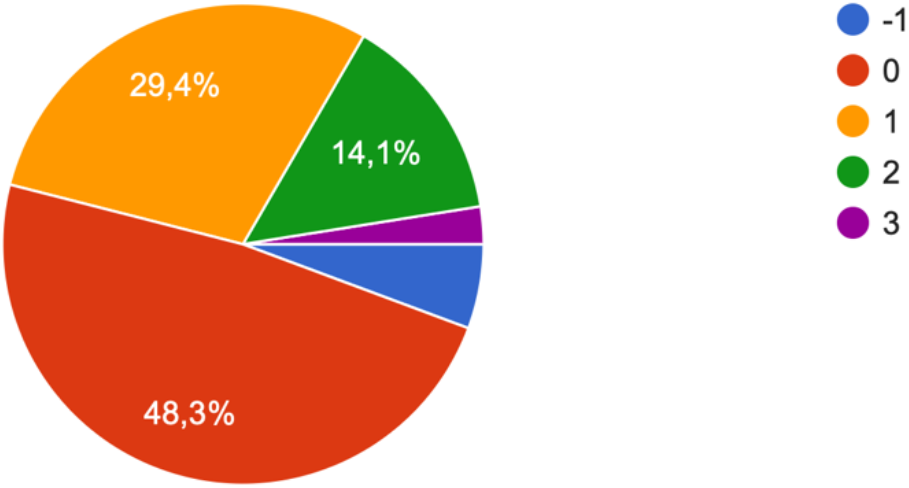

Analyzing the ***symptom complex reflecting the deterioration of the cardiovascular functional system*** in Ukrainian youth, the following features were established.

4.4% of respondents noted that this feature was characteristic of them before. 63.4% of respondents practically do not feel the deterioration of the cardiovascular functional system,

23.9% of respondents have certain signs of deterioration of the cardiovascular functional system, but they do not significantly affect life and well-being.

At the same time, 7.1% of respondents noted that the manifestation of this symptom complex is strong, and 1.1% feel exhausted and disorganized due to the deterioration of the cardiovascular functional system.

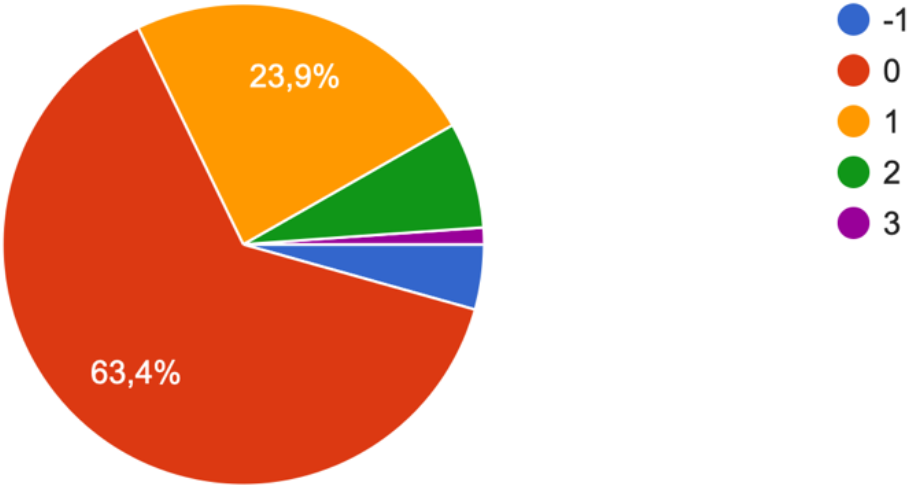

Regarding ***the symptom complex reflecting the deterioration of the functional hematopoietic system (anemic syndrome)***, the respondents evaluated their experience in the following way.

4.5% of respondents used to have this symptom, but now it has disappeared or does not bother them. 78.4% do not experience these symptoms. 12.1% of respondents 1 have certain signs of anemic syndrome, but they do not significantly affect life and well-being. However, 4.1% of respondents say that the manifestation of this feature is strong, and only 0.9% feel exhausted and disorganized.

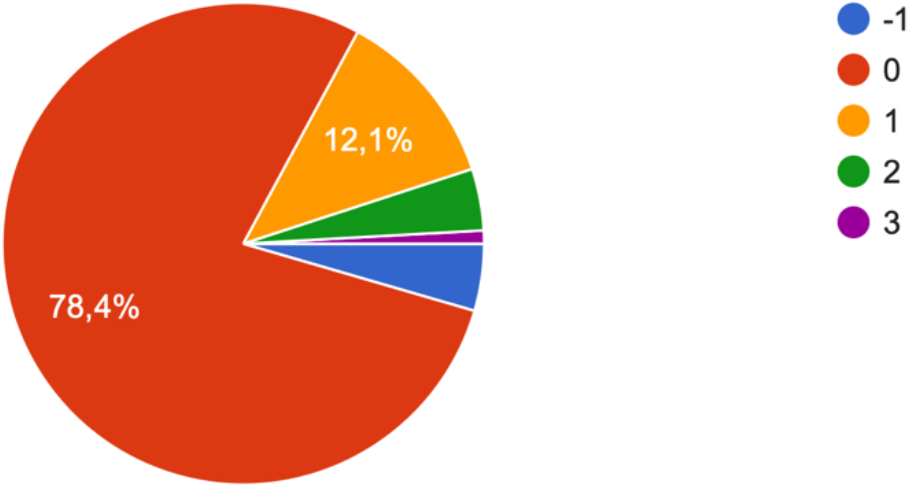

According to the results of the self-assessment of the ***symptom complex, which reflects the deterioration of the immune functional system (allergies)***, the respondents gave the following answers.

5.1% of respondents indicate that this feature used to be characteristic of them, but now it has disappeared or does not bother them. At the same time, 64.7% claim that they practically do not feel the deterioration of the immune functional system, while 20.4% of respondents have certain signs of it, but they do not significantly affect life and well-being. It is noteworthy that 8.2% of respondents have a strong manifestation of this symptom, and 1.6% feel exhausted and disorganized.

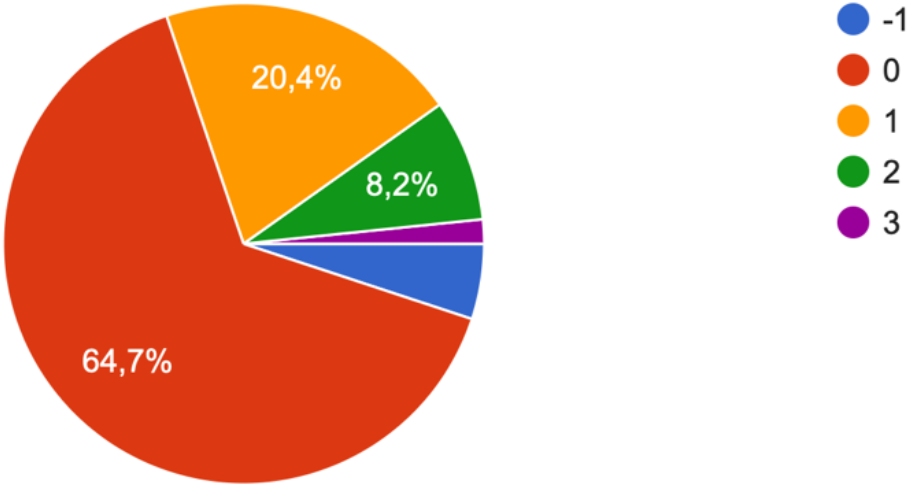

Signs of ***vascular dysregulation (water balance, thermoregulation, sweating, etc.), which, as a rule, is formed under the influence of traumatic mental factors*** in respondents have the following manifestations.

3.5% noted that this symptom complex used to be characteristic of them, but now it has disappeared or does not bother them. 63.5% do not experience these symptoms or they are absent. 23.4% of respondents have certain signs of vascular dysregulation, but they do not significantly affect life and well-being.

At the same time, 8.1% have a strong manifestation of this symptom, and 1.5% feel exhausted and disorganized.

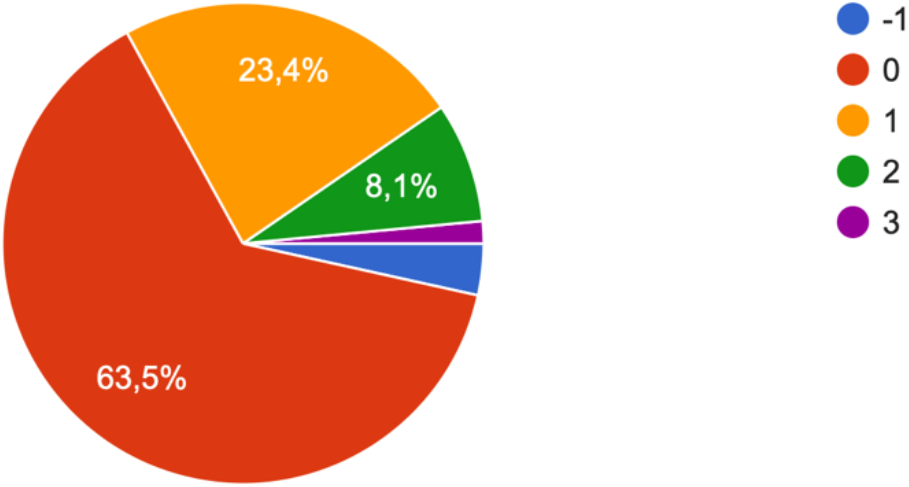

Regarding the ***problems of physical functioning*** of Ukrainian youth in war conditions, the following was established.

3.5% of respondents experienced physical functioning problems before, but now this problem does not bother them. 63.6% of respondents do not experience physical functioning problems. 23.6% have certain problems, but they do not significantly affect their life and well-being.

At the same time, for 7.4% of respondents, the manifestation of this symptom is strong, and 1.9% feel exhausted and disorganized.

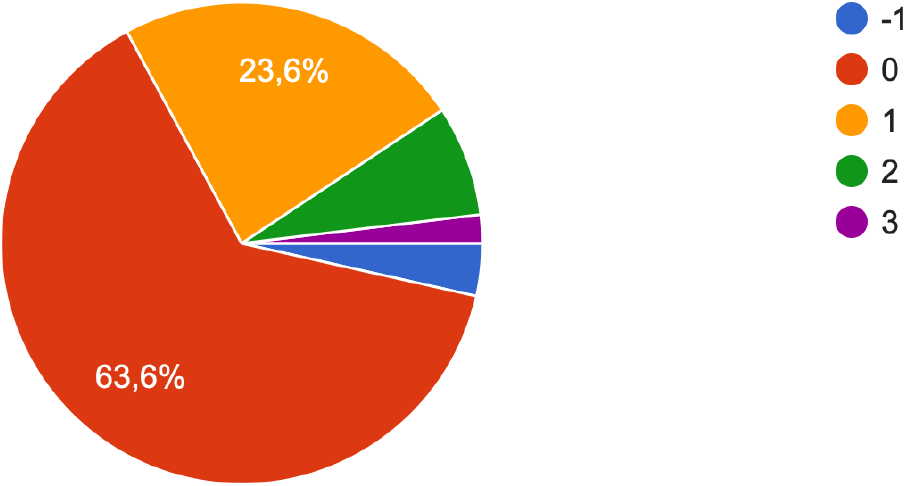

Assessing their own ***role functioning problems caused by physical condition***, the respondents noted the following.

Thus, 2.7% of respondents note that they have gotten rid of the influence of physical condition on role functioning. Similarly, 77.8% of respondents note that they do not feel the corresponding influence.

At the same time, 15.1% of Ukrainian youth say that there are certain signs of the problem of role functioning, which are caused by physical condition, but they do not significantly affect life and well-being. In 3.6%, problems of role functioning are quite strong, and 0.8% feel exhausted and disorganized).

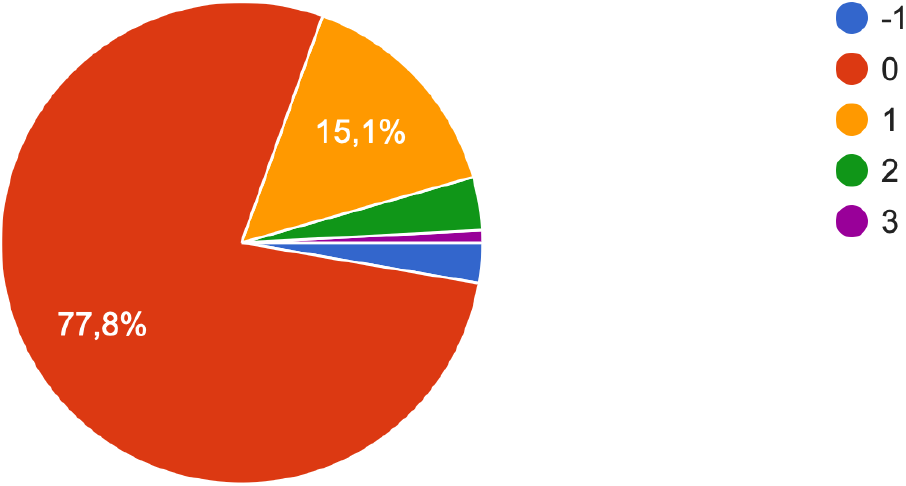

The next question for self-assessment was the “***Intensity of pain***” indicator.

As determined by 4.7% of respondents, the intensity of physical pain was characteristic of them earlier, but now it has disappeared or does not bother them. 61.3% of respondents now practically do not feel physical pain, or it is absent. However, 25.1% of respondents have certain signs of feeling physical pain, but they do not significantly affect their life and well-being.

At the same time, 7.6% of respondents note that the manifestation of physical pain is strong, and 1.3% of respondents feel exhausted and disorganized due to physical pain.

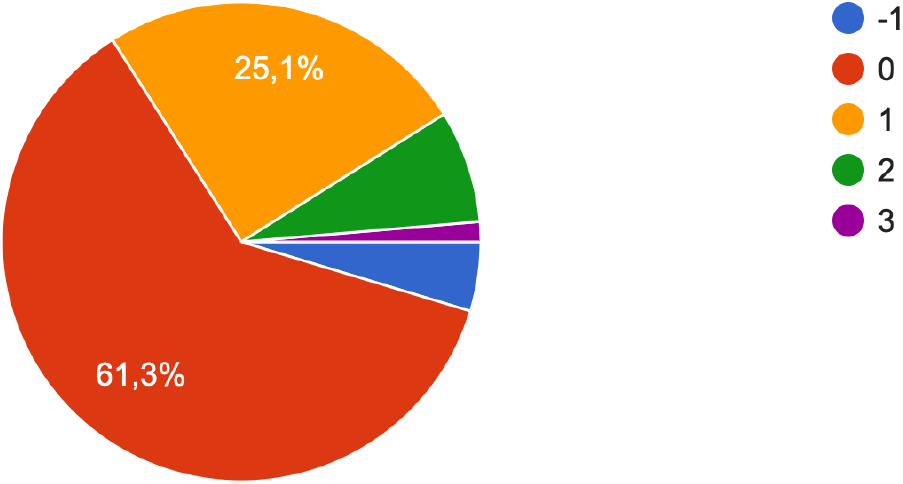

The respondents’ self-assessment of the “***Life activity***” indicator looks as follows.

9.4% of respondents believe that they used to be characterized by life activity, but now it has disappeared. 21.1% of respondents practically do not feel their characteristic level of vital activity.

At the same time, 37.8% of respondents believe that the level of life activity does not significantly affect their life and well-being. At the same time, 23.4% of respondents claim a high level of vitality, and 8.3% of respondents feel exhausted and disorganized.

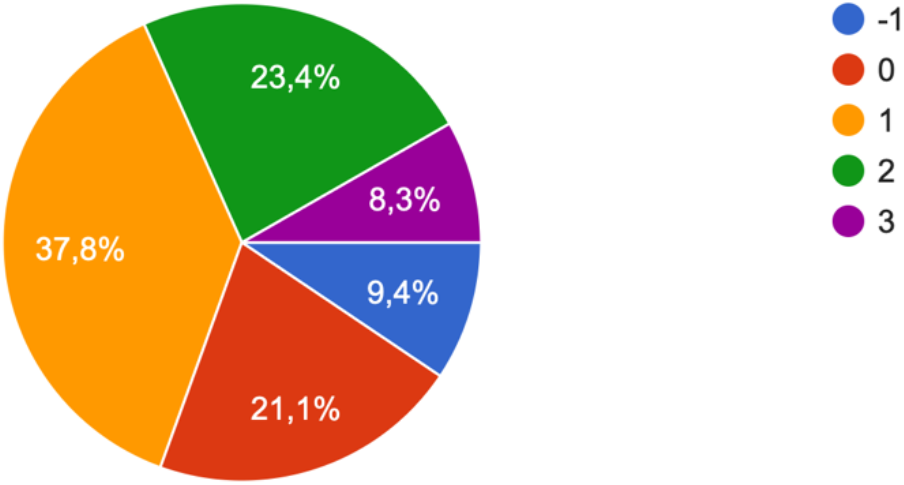

The respondents noted the following about the ***problems of social functioning***.

5.6% of respondents noted that they had problems with social functioning before, but now they have disappeared or do not bother them. 42.0% of respondents do not experience problems with social functioning. 31.4% of respondents say that there are certain signs of problems with social functioning, but they do not significantly affect life and well-being.

At the same time, 16.1% of respondents say that the manifestation of this feature is strong, and 5.0% of respondents feel exhausted and disorganized due to problems with social functioning.

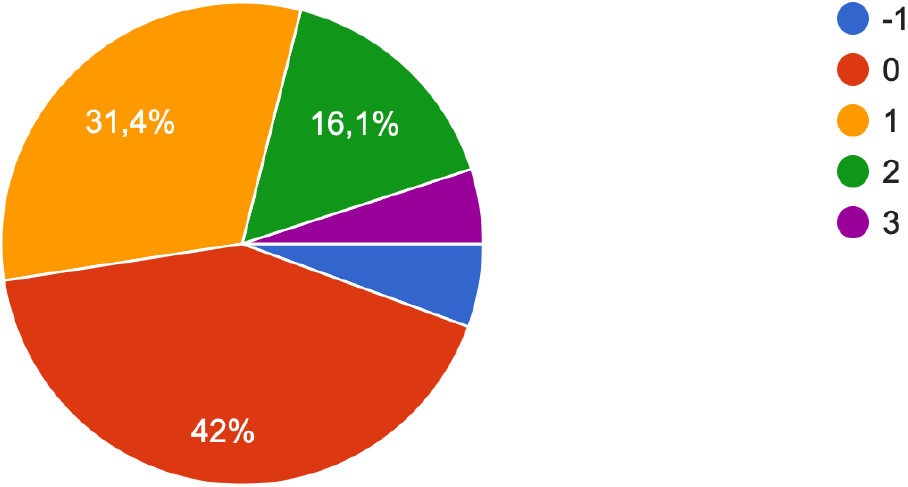

The respondents’ self-assessment of ***the problems of their own role functioning, caused by the emotional state, l***ooks like this distribution.

2.6% of respondents say that earlier this feature was characteristic of them, but now it has disappeared or does not bother them. 49.9% of respondents practically do not experience problems of their own role functioning caused by emotional state. 30.0% of respondents determined that they have certain signs of role functioning problems, but they do not significantly affect life and well-being.

In addition, 13.7% of respondents note that the manifestation of this symptom is strong, and 3.8% of respondents have a feeling of exhaustion and disorganization due to problems in their own role functioning caused by their emotional state.

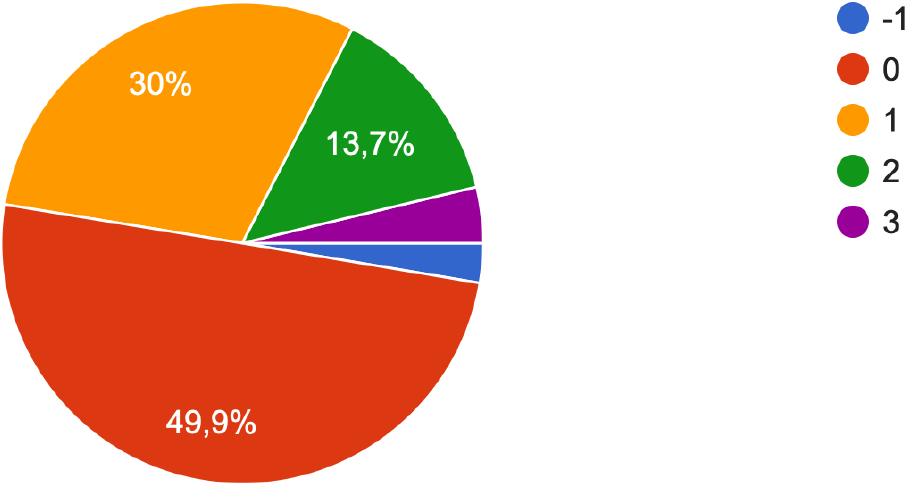

When giving a self-assessment of ***problems related to the mental health***, the respondents noted the following.

3.9% of respondents noted that problems related to mental health previously were characteristic of them, but now they have disappeared or do not bother them. 32.8% of young people practically do not experience such problems. 34.5% of respondents say that they have certain problems with their mental health, but they do not significantly affect their life and well-being.

At the same time, which requires the attention of psychologists, 20.8% of respondents say that the manifestation of this sign is strong, and 8.0% of the interviewed youth feel exhausted and disorganized in connection with the state of mental health.

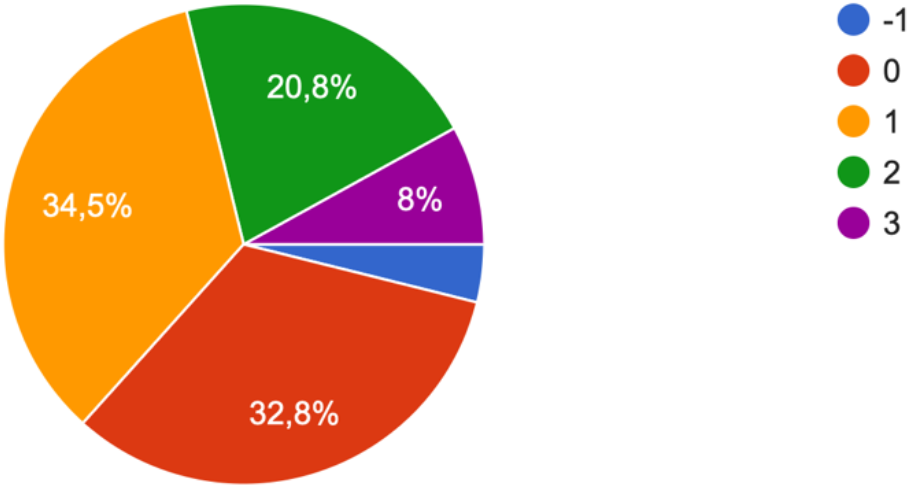

### Behavioral changes in young people that notice in everyday behavior and activity

The next block of research questions was related to behavioral changes in young people that they themselves and their close people notice in everyday behavior and activity. The obtained results allow us to imagine the behavioral portrait of Ukrainian youth in everyday life.

When answering the question “***I and/or my loved ones notice that my appearance has changed”***, the respondents noted the following.

2.4% of respondents noted that changes in their appearance used to be characteristic of them, but now such changes do not occur. 46.1% of respondents do not show changes in appearance.

However, 31.4% of respondents have certain signs of changes in appearance, but they do not significantly affect life and well-being.

At the same time, 16.6% of surveyed youth notice strong changes in appearance, and 3.5% of respondents feel exhausted and disorganized due to how their appearance has changed.

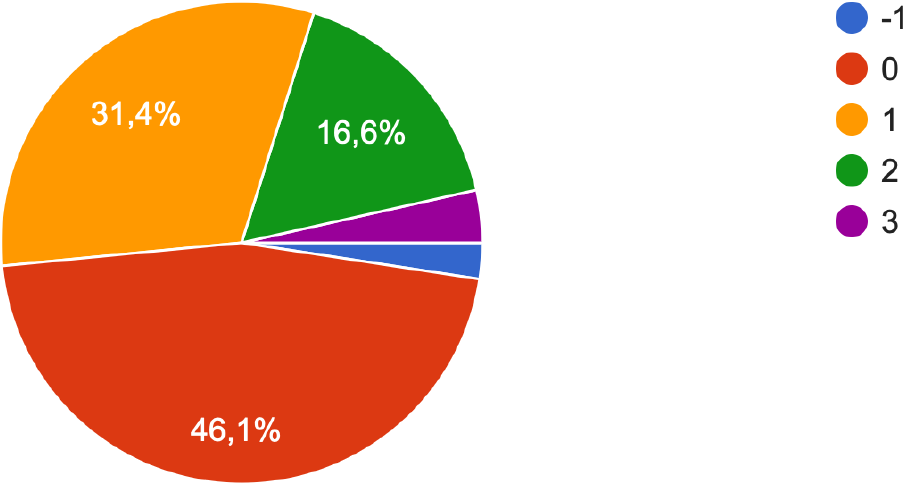

Regarding the question “***I and/or my loved ones notice that my gait has changed***”, the following answers were received from the respondents.

3.5% of respondents previously observed similar changes in themselves, but currently no such changes are occurring. 86.4% of respondents say there are no such changes.

At the same time, 6.9% of respondents notice certain signs of change in gait, but they do not significantly affect life and well-being. 2.8% of respondents say that the manifestation of such changes is strong, and 0.5% of respondents feel tired and disorganized due to the change in gait.

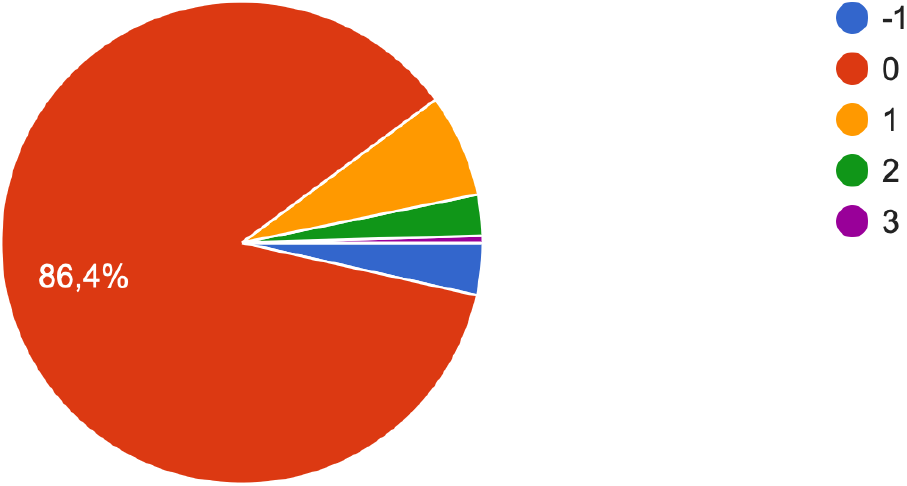

Evaluating the question “***I and/or my loved ones notice that my way of dressing has changed***”, the young people interviewed note the following.

2.7% of respondents note that they used to change their way of dressing, but now this tendency has disappeared. 62.2% of respondents did not change the way they dress. 22.3% of respondents experienced such changes, but they do not significantly affect life and well-being.

At the same time, 10.5% of respondents say about significant changes in the way they dress, and 2.2% of young people claim that they feel exhausted and disorganized due to changes in the way they dress.

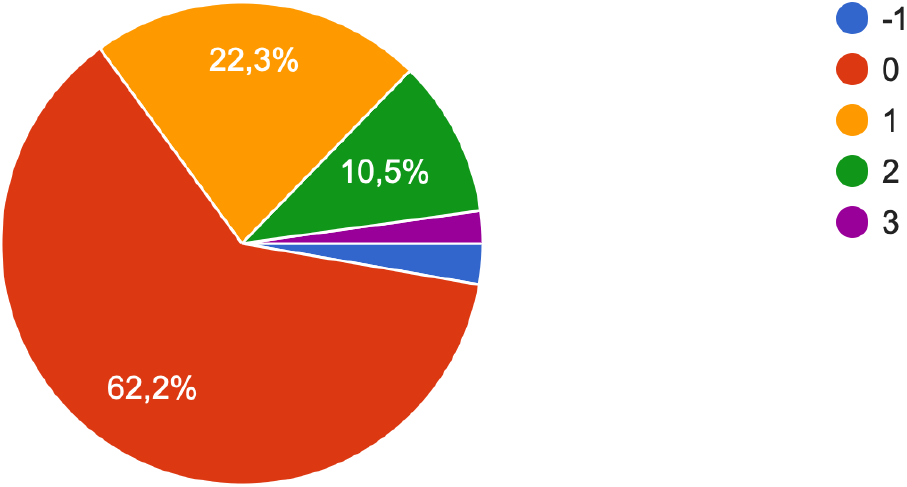

Regarding the question “***I and/or my loved ones notice that my posture has changed***», the respondents answered the following.

3.1% of respondents used to have this feature, but now it has disappeared. 80.8% of respondents do not notice changes in their posture. However, 11.2% of respondents note that there are certain changes in their posture, but they do not significantly affect their life and well-being.

4.2% of respondents were forced to change their posture in new conditions, and 0.7% of respondents feel exhausted and disorganized due to such changes.

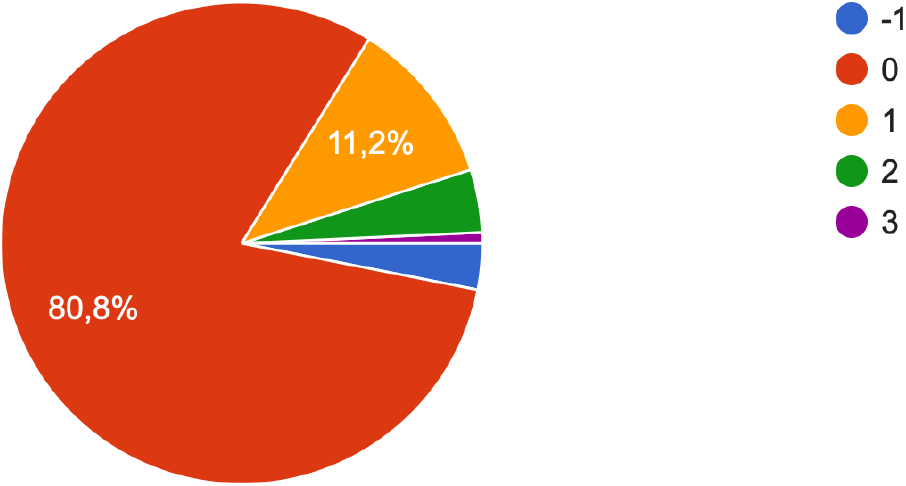

It should be noted that when answering the question “***I and/or my loved ones notice that my body movements have changed***”, the respondents noted the following.

2.9% of respondents note that they used to have occasional changes in body movements, but now they do not notice such changes. 79.2% of respondents do not experience changes in body movements.

At the same time, 13.4% of respondents notice certain changes in body movements, but they do not significantly affect life and well-being. At the same time, 3.9% of the surveyed youth say that the manifestation of this symptom is strong, and 0.7% feel exhausted and disorganized) due to changes in body movements.

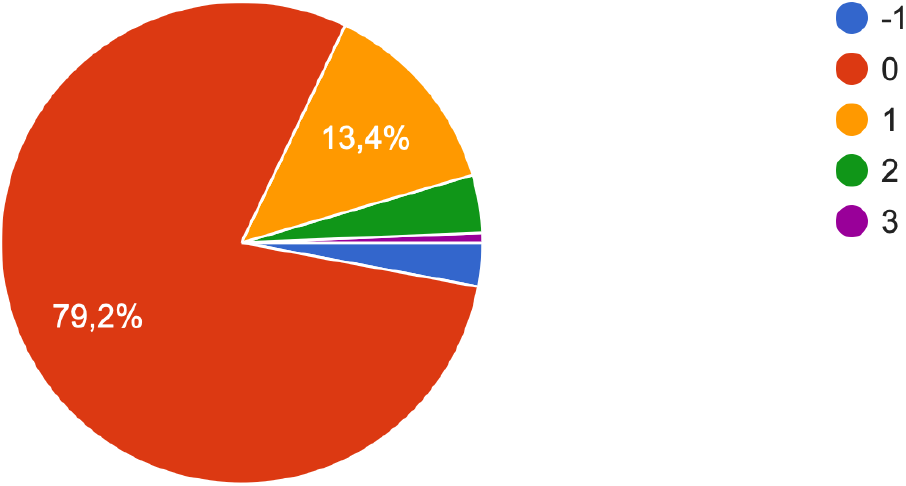

Regarding the question “***I and/or my loved ones notice that my behavior has changed***”, the following results were obtained.

1.4% of respondents note that they used to experience changes in their behavior, but now such changes do not occur. 32.5% of respondents do not feel changes in their behavior, or they are absent.

At the same time, 39.8% of respondents confidently testify that there are certain signs of behavior change, but they do not significantly affect life and well-being.

On the other hand, 22.3% of respondents testify that the manifestation of changes in behavior is strong, and 4.1% feel exhausted and disorganized due to the change in their own behavior.

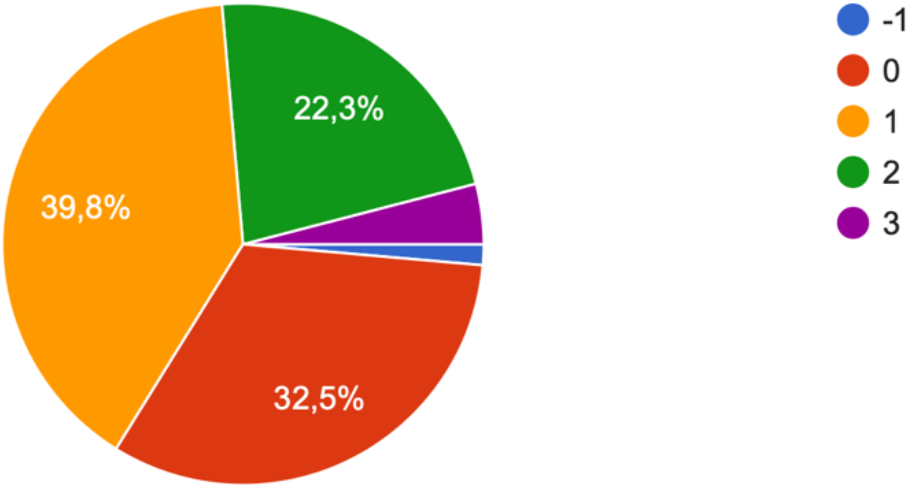

When giving a self-assessment of the change in behavior to the question “***I and/or my loved ones notice that my speech has changed in tempo***”, the respondents noted the following.

Thus, 2.4% of respondents indicate that this feature used to be characteristic of them, but now it has disappeared or does not bother them. 70.1% of respondents practically do not feel changes in speech in terms of tempo.

At the same time, 18.4% of the surveyed youth say that there are certain signs of a change in speech tempo, but they do not significantly affect life and well-being.

However, for 7.6% of respondents, the manifestation of these changes is strong, and 1.5% of young people feel exhausted and disorganized because the pace of their speech has changed.

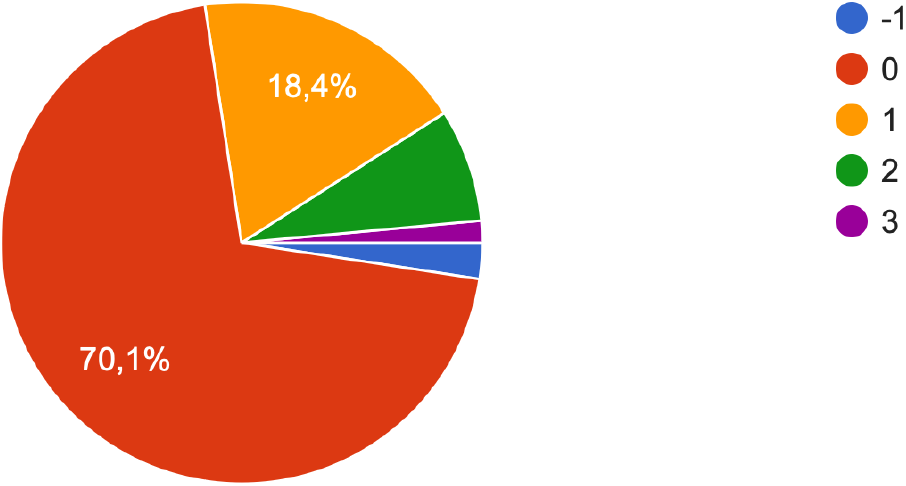

The following results were obtained regarding the question “***I and/or my loved ones notice that my speech has changed in terms of emotional coloring***.”

1.9% of the interviewed youth say that earlier this feature was characteristic of them, but now it has disappeared or does not bother them. 50.1% of respondents say that they have no changes in the emotional color of their speech. In addition, 30.8% show certain changes in the emotional coloring of speech, but they do not significantly affect life and well-being.

At the same time, 14.2% of the respondents have a strong manifestation of this change, and 3.0% feel exhausted and disorganized.

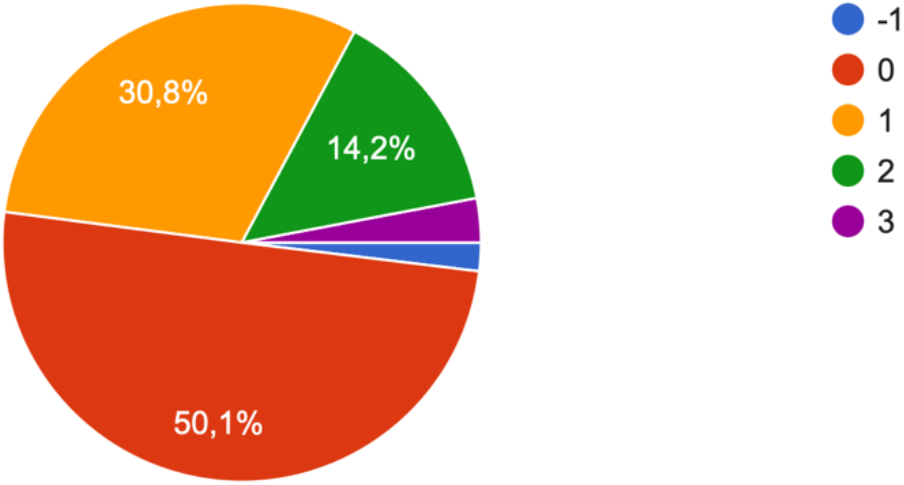

About the statement “***I and/or my loved ones notice that my speech has changed logically***”, such a distribution was found among the surveyed youth.

2.5% of respondents note that this feature used to be characteristic of them, but now it has disappeared or does not bother them. 67.1% of respondents claim that they practically do not feel such changes. But 20.0% of the interviewed youth have certain signs of changing the logic of speech, but they do not significantly affect their life and well-being.

9.1% of respondents notice that the manifestation of this sign is strong. At the same time, 1.3% feel exhausted and disorganized due to changes in the logic of speech.

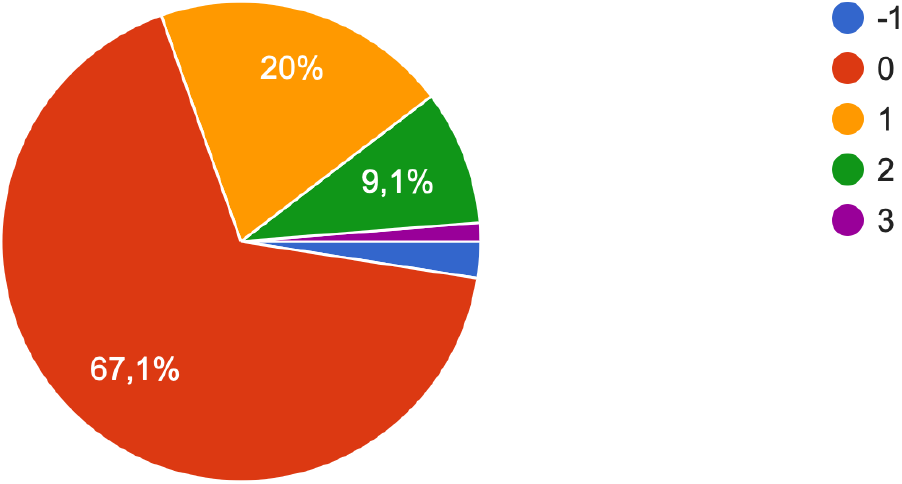

Finally, evaluating the statement “***I and/or my loved ones notice that my autonomic reactions have changed***”, the following results were obtained.

2.3% of the interviewed youth believe that this feature used to be characteristic of them, but now it has disappeared or does not bother them. 77.6% of young people practically do not experience changes in vegetative reactions. However, 15.8% of respondents have certain signs of changes in vegetative reactions, but they do not significantly affect life and well-being.

3.6% of respondents reported strong changes in autonomic reactions, and 0.7% of respondents felt exhausted and disorganized due to autonomic changes.

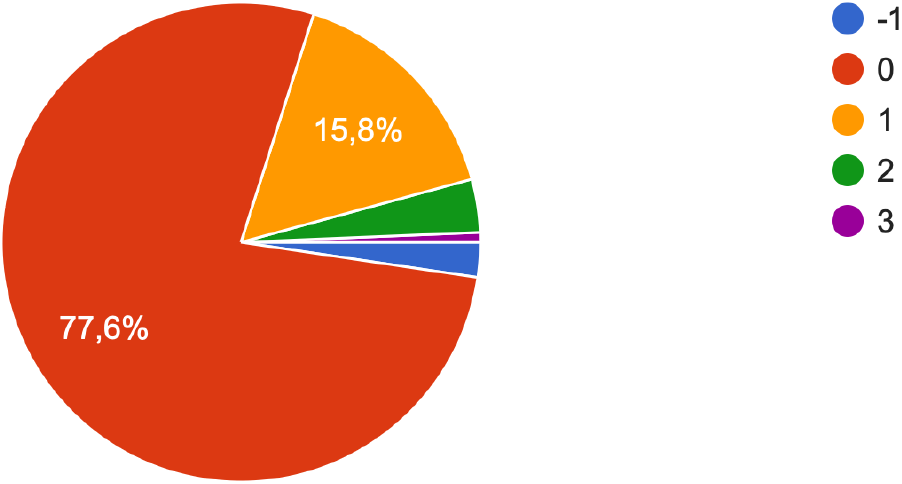

## Conclusions

Such awareness becomes the basis for understanding the new norm of life during the war.

## Data Availability

All data produced in the present study are available upon reasonable request to the authors

https://forms.gle/U99akzToALh2mXLc9

## Declaration of Conflicting Interests

The author(s) declared no potential conflicts of interest with respect to the research, authorship, and/or publication of this article.

**This study did not receive any funding**

**Ethics committee of G.S. Kostyuk Institute of Psychology of the National Academy of Educational Sciences and Department of General and Medical Psychology of the Bogomolets National Medical University of Ukraine (protocol No. 15 dated 31.03.2022) gave ethical approvals for this work**.

## Notes

### Competing Interest Statement

The authors have declared no competing interest.

### Author Declarations

Ethics committee of G.S. Kostyuk Institute of Psychology of the National Academy of Educational Sciences and Department of General and Medical Psychology of the Bogomolets National Medical University of Ukraine (protocol No. 15 dated 31.03.2022) gave ethical approvals for this work.

